# In the long shadow of our best intentions: model-based assessment of the consequences of school reopening during the COVID-19 pandemic

**DOI:** 10.1101/2020.09.18.20197400

**Authors:** Kaitlyn E. Johnson, Madison Stoddard, Ryan P. Nolan, Douglas E. White, Natasha S. Hochberg, Arijit Chakravarty

## Abstract

As the United States grapples with the ongoing COVID-19 pandemic, a particularly thorny set of questions surrounds the reopening of K-12 schools and universities. The benefits of in-person learning are numerous, in terms of education quality, mental health, emotional well-being, equity and access to food and shelter. Early reports suggested that children might have reduced susceptibility to COVID-19, and children have been shown to experience fewer complications than older adults. Over the past few months, our understanding of COVID-19 has been further shaped by emerging data, and it looks increasingly likely that children are as susceptible to infection as adults and have a similar viral load during infection. While the higher prevalence of asymptomatic disease among children makes symptom-based isolation strategies ineffective, asymptomatic patients do not in fact carry a reduced viral load. Using assumptions consistent with the emerging understanding of the disease, we conducted epidemiological modeling to explore the feasibility and consequences of school reopening in the face of differing rates of COVID-19 prevalence and transmission. Our findings indicate that, regardless of the initial prevalence of the disease, and in the absence of systematic surveillance testing, most schools in the United States can expect 20-60 days without a major cluster emerging. Without testing or contact tracing, the true extent of these disease clusters may not be apparent, and our research suggests that the case count will underestimate the true size of the clusters by a large margin. These disease clusters, in turn, can be expected to propagate silently through the community, with potentially hundreds to thousands of additional cases resulting from each individual school cluster. Thus, our findings suggest that the debate between the risks to student safety and benefits of in-person learning frames a false dual choice. Given the current circumstances in the United States, the most likely outcome in the late fall is that students will be deprived of the benefits of in-person learning while having incurred a significant risk to themselves and their communities.

## Introduction

As is to be expected with any emerging infectious disease, our understanding of the biology and transmission of COVID-19 continues to evolve rapidly during this ongoing pandemic. In particular, changes in our understanding of the disease impact our expectations of the risk to children and the community that would arise from the reopening of K-12 schools and colleges.

A number of studies at the outset of the pandemic suggested that children were less susceptible, with a lower risk of being infected with COVID-19 upon exposure to the virus (Viner *et al*., 2020). Children and young adults in general were also found to have mild symptoms of the disease, with low rates of hospitalization and death (CDC COVID-19 Response Team, 2020; Isaacs and Hos-, 2020; Song *et al*., 2020). For this demographic group, asymptomatic and pauci-symptomatic cases were also observed at a high frequency (CDC COVID-19 Response Team, 2020; Chen *et al*., 2020; Dong *et al*., 2020; Qiu *et al*., 2020; Wu and McGoogan, 2020). Some early reports also suggested a lower rate of infection (attack rate) in children (Dimeglio *et al*., 2020; Jing *et al*., 2020; Mizumoto, Omori and Nishiura, 2020). However, these findings were confounded with widespread school closures (Jing *et al*., 2020) and the potential for a bias in testing due to undercounting the asymptomatic cases. These findings were also contradicted by other reports suggesting no difference in attack rates between children and adults (Fontanet *et al*., 2020). A key finding reported and cited often in the early debate about school reopening was that children were not usually the index case (first infection) within in a family (Zhu *et al*., 2020), suggesting that children may not be responsible for disease spread (Couzin-Frankel, Vogel and Weiland, 2020; Viner *et al*., 2020).

Based on this scientific understanding at the time, and mindful of the harm to children’s long term development in the face of prolonged school closures, a number of medical associations and public health figures strongly advocated for a return to in-person schooling (American Academy of Pediatrics, 2020; Munro and Faust, 2020; National Association of School Nurses, 2020; The National Academy of Sciences Engineering and Medicine, 2020) even going so far as to endorse a return to in-person schooling with closer spacing than recommended by the CDC (American Academy of Pediatrics, 2020). A lengthy white paper on the CDC’s own website also argues this point, heavily emphasizing the harm to children that results from loss of in-person educational instruction and school resources (CDC, 2020c). The harm to children’s development, to their psychological well-being (particularly for teenagers), the potential risks to vulnerable children, and the increase in inequality that results from school closure is well documented (Auxier and Anderson, 2020; Fitzpatrick *et al*., 2020; Loades *et al*., 2020) and frames a strong case for a return to in-person schooling if the biology of COVID-19 supports it.

In recent months, our fundamental understanding of the disease has shifted under our feet. First, a number of studies demonstrated that children’s susceptibility to COVID-19 is similar to that of adults (Bi *et al*., 2020; ONS, 2020; Zhang *et al*., 2020), and there have been numerous publicized examples of peer-to-peer spread among children in congregate settings, particularly without masks (Stein-Zamir *et al*., 2020; Szablewski *et al*., 2020). These findings suggest that the low attack rate observed in children during the early days of the pandemic may have been a function of school closures and other behavioral changes (Jing *et al*., 2020) rather than reduced susceptibility among children. Second, viral loads in children have been found to be similar or arguably higher than those of severely ill adults (Heald-Sargent *et al*., 2020; Jones *et al*., 2020; Yonker *et al*., 2020) with prolonged fecal shedding a particular feature of the disease course (Xu *et al*., 2020). Third, and most tellingly, a large proportion of COVID-19 cases in children and young adults has been found to be asymptomatic (Bi *et al*., 2020; DeBiasi and Delaney, 2020; Sola *et al*., 2020). This last finding casts further doubt on the early reports of lower attack rates and transmission from children, as asymptomatic cases were often missed in epidemiological tracing studies during the early stages of the pandemic.

Other aspects of our understanding of COVID-19 spread have also evolved over time. In the early days of the pandemic, guidelines for preventing the spread of the disease were heavily focused on respiratory droplets and transmission from fomites (objects contaminated with the virus). As our understanding of the disease has matured, fomites have been recognized to be less of a threat (Goldman, 2020; WHO, 2020). On the other hand, airborne transmission via small aerosolized droplets has been identified as a plausible route of disease spread (Jayaweera *et al*., 2020; Morawska and Milton, 2020). There are multiple documented cases of indoor spread that can best be explained by airborne transmission (for a summary, see (Qureshi *et al*., 2020), and Supplementary Table S4). First-principles calculations of viral load and droplet physics add further credence to the view that transmission via small aerosolized droplets represents a tangible threat in indoor environments (Basu, 2020), a view that is shared by the WHO (WHO, 2020) and CDC (CDC, 2020a). Consistent with this, COVID-19 is difficult to control in indoor settings, and spread can occur over short time periods, even in the presence of extreme precautions (Bae *et al*., 2020). One report based on contact tracing of clusters of cases occurring in Japan estimated the odds of transmission in an enclosed environment to be 18.7 fold higher than in an outdoor environment (Nishiura *et al*., 2020), at a time (February 2020) when mask-wearing in Japan may have been generally prevalent (Kyodo News, 2020).

While our understanding of SARS-CoV-2 transmission has altered rapidly over the past few months, guidance for infection prevention has not kept pace. The CDC briefly acknowledged that the disease can be spread in poorly ventilated indoor spaces (CDC, 2020a; The New York Times, 2020b), even at distances greater than six feet. However, their guidance continues to emphasize the six-foot rule, which was originally intended to minimize exposure to cough and sneeze trajectories. Thus the guidance is not necessarily adequate to limit SARS-CoV-2 spread (Qureshi *et al*., 2020). Guidance also strongly emphasize handwashing and surface cleaning, even though the CDC’s recent statements have emphasized that surface contamination is not a main driver of SARS-CoV-2 spread (CDC, 2020a). Many schools are still following the original CDC guidelines from the early days of the disease (CDC, 2020b, 2020d). To the extent that a gap may or may not have opened up between the CDC’s understanding of the disease (CDC, 2020a) and the official guidelines for school reopening (CDC, 2020d), the effectiveness of these guidelines-even for schools with the resources to follow them perfectly-is unknown.

Taken together, this state of affairs raises the possibility that children may be a potential source of contagion for COVID-19, and schools -despite following guidelines-may be inadequately prepared for this threat. An outbreak seeded among children may in fact result in transmission chains that are harder to bring under control, as the asymptomatic nature and milder presentation of infected children will delay the detection of the disease in the absence of widespread, rapid, molecular testing.

With this in mind, we have conducted a model-based investigation of the feasibility and consequences of school reopening within the United States during the SARS-CoV-2 pandemic.

## Methods

### SEIR model of SARS-CoV-2

Our analysis was based on a standard susceptible-exposed-infected recovered (SEIR) epidemiological model (Li and Muldowney, 1995). We assumed an equivalent level of infectivity and susceptibility between children and adults, a conservative assumption in the face of our updated understanding of the situation in children for the current pandemic. Susceptible individuals (S) can become infected at a rate proportional to the number of infected individuals. We assumed a constant infectiousness throughout the duration of infection, as is standard for SEIR models. Although infectiousness is expected to vary over time, we did not explicitly account for this. In a previous study we showed that given the kinetics of SARS-CoV-2 infectiousness, isolating subsequent to development of symptoms has minimal impact on reduction of disease spread (Johnson *et al*., 2020).

Susceptible individuals (S) become exposed (E) at a rate (β) proportional to the number of individuals infected (I). Infected individuals were assumed to remain exposed but not detectable or infectious (E) for an average of 3 days, corresponding to a rate α (Lauer *et al*., 2020). A proportion of the infected individuals (I) develop symptoms after an average of 2.3 days of being infectious (He *et al*., 2020). Infectious individuals recover after an average of 14 days, corresponding to a rate of γ (He *et al*., 2020). The model equations are as follows:

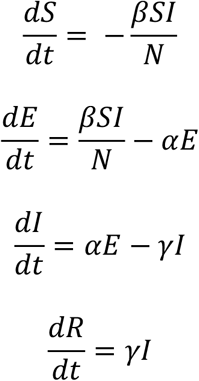

Where *N* = total size of the school population. The model assumes that the students are a well-mixed population, and therefore does not take into account any cohorting or podding. The model does not explicitly account for reduced transmission on the weekends, and so all references to days corresponds to total epidemic days, not school days. The initial number of infected individuals in a school is varied based on the community prevalence that is simulated. For communities with a prevalence expected to be lower than 1 in 1000, we assumed the population was sufficiently large and well-mixed that it was appropriate to assume a deterministic solution (Rouzine, Rodrigo and Coffin, 2001) with an initial condition of *Np* where *p* is the prevalence per 1000 in that community. Model parameters and their references are described in more detail in Table S1.

### Estimation of time to detection of first case

To simulate the expected detected cases over time, we transformed the cumulative infections over time from the infected compartment of the SEIR model in the following ways. First, we assumed cases are only detected via symptomatic students seeking testing after symptom onset. Proactive surveillance testing at schools was not assumed, as this is not part of CDC guidelines for school reopening (CDC, 2020d), and many public K-12 schools in the country are not currently performing this. We assumed students develop symptoms in only 21% of cases (Davies *et al*., 2020). In the infectious individuals that eventually develop symptoms, the time to detect cases is delayed. We assumed a 2.3 day delay from infectiousness to symptom onset, and a further optimistic 0.7 day delay until seeking a test. Then we assumed the delay to receive test results was an additional 4 days, consistent with the national average (Baum *et al*., 2020). Based on our recent analysis (Johnson *et al*., 2020) demonstrating that a delay of more than a day from symptom onset to isolation renders the strategy minimally effective at reducing disease transmission, we did not explicitly account for removal of symptomatic students in this model. We report the expected time to the first detected case and the number of cumulative true infections at that time (Supp. Fig. S1) to estimate the magnitude of latent infections in the school when the first case is detected.

### Estimation of time to close

To simulate a school closing, we had to make some assumptions about the “tolerance” of a school district. Based on recent university closings (Redden, 2020; Richard Fausset, 2020; Snyder, 2020b), we reasoned that a cumulative incidence of detected cases exceeding 1% of the student body would initiate moving schools online. This corresponds to 10 students in a school of 1000. We reasoned that each student had on average 10 contacts and report the percent quarantined over time (Supp. Fig. S2). We note that this does not account explicitly for any podding or cohorting measures in place that may violate the well-mixed population assumption and prevent the need to close all sections of the school population. We report the number of cumulative student infections at the time that the school closes.

### Estimation of effect of secondary spread within the community

Because student infections pose a significant risk of infecting their family members and thus the surrounding community, we performed an analysis to assess what this risk might be in terms of secondary community infections. Since school outbreaks are expected to occur fairly quickly, we treated the number of infections at school closure as the initial condition for a community epidemic and used an SEIR model to estimate the scale of this secondary cluster. For a given community R_0_ and number of initially seeded student infections, we then modeled the number of cumulative additional infections expected within the community in 100 days after the infections were seeded by the school. The presence of immune individuals within the community is implicitly incorporated into the low community R_0_, which may be due to a combination of immunity and social distancing measures.

### Estimation of average prevalence

The average initial prevalence of infection in students is assumed to be equal to that of their community. Community prevalence was chosen based on the calculated prevalence of a number of counties in the United States. To calculate the prevalence of a county, we used the New York Times database of daily cases per 100k by county. We assumed an infectious duration of 7 days and a reporting rate of 1 in 5 (with scenario lower and upper bounds of 1 in 3 and 1 in 10). We assumed the initial students that would arrive infected in the first week in a school of 1000 would be sampled evenly from this county prevalence (Glanz, Carey and Cohen, 2020). A table of county prevalence values for some representative regions in the US are shown in Table S2, and an example of how to estimate these from the New York Times daily case counts per 100k in each county is demonstrated in Table S2. For simulations varying the effect of R_0_ on infections, we assumed a baseline prevalence close to the national prevalence of 5 in 1000 as of August 27th, 2020 (The New York Times, 2020a) with upper and lower scenario bounds of 10 in 1000 and 3 in 1000 respectively.

### Estimation of school R_0_

It is not well-known empirically what a school’s reproductive number is at baseline due to the lack of early epidemic data. We reasoned that schools would vary significantly in their ability to reduce transmission, partially as a function of their resources. As of this writing, there have been multiple documented cases of schools across the country reopening without providing adequate personal protective equipment (PPE), mandating mask use or enforcing physical distancing (Andone and Johnston, 2020; CBS 4 New York, 2020; Tutman and Dado, 2020). On the other hand, some schools have gone beyond the CDC guidance, implementing hybrid schooling, upgrading their ventilation systems, mandating masks and providing adequate PPE to their students and staff. Modeling the impact of specific interventions on the R_0_ is beyond the scope of this paper, and we have instead varied the R_0_ over a wide range, focusing our attention on the downstream consequences of reopening given a particular R_0_.

We reasoned that an indoor setting, where masks and distancing might vary in adherence, could have a range of R_0_ values from as low as just above 1 to as high as 5. (Some estimates of the R_0_ of SARS-CoV-2 in different settings are presented in Supplementary Table S3 for comparison, and some examples of evidence supporting efficient indoor transmission are provided in Supplementary Table S4). For simulations varying the initial prevalence, we assumed a baseline R_0_ of 2.5, with upper and lower scenario bounds of 3.5 and 2.2 respectively.

Unfortunately, it is unlikely that schools will have any knowledge of their R_0_ prior to reopening. Thus, it is our suggestion that the reader considers the outcomes at both the low and the high end of the R_0_ range as being within the realm of the possible for their own situation.

## Results

### Clusters will develop quickly in schools upon reopening

Our analysis suggests that most schools in the United States will experience outbreaks within a short time of reopening (Figure 1) for a wide range of initial disease prevalences and school R_0_. Focusing on Figure 1A & B, where school R_0_ was held constant at 2.5, we draw the reader’s attention to the line representing a prevalence of 1 in 1000. This prevalence, which a number of localities have set as their “green zone” guideline for lowest concern (for example (Gentile, 2020)), only results in a delay of 45 epidemic days until the outbreak reaches a detectable size to prompt school closures (we are optimistically assuming that 1% of the student body having a detected case is sufficient to trigger closing). We note that a number of schools have already shut down at the first detection of cases (Hanau, 2020), or will have to shut down before this threshold is reached due to illness in staff and/or absenteeism due to quarantine/fear. Of course, the impact of detected case load on school closing decisions is impossible to model. However, somewhat obviously, if a school chooses to remain open in the face of continued spread of the virus (without implementing aggressive measures to control spread, such as contact-tracing based isolation), it can expect about 80% of the students infected regardless of initial prevalence. It is worth noting that the national average prevalence of active infections of COVID-19 within the United States, at the time of this writing, is 5/1000, which corresponds to the yellow line in Figure 1A. At this prevalence, schools can expect to remain open for about 4 weeks, given the 1% threshold.

**Figure 1:**
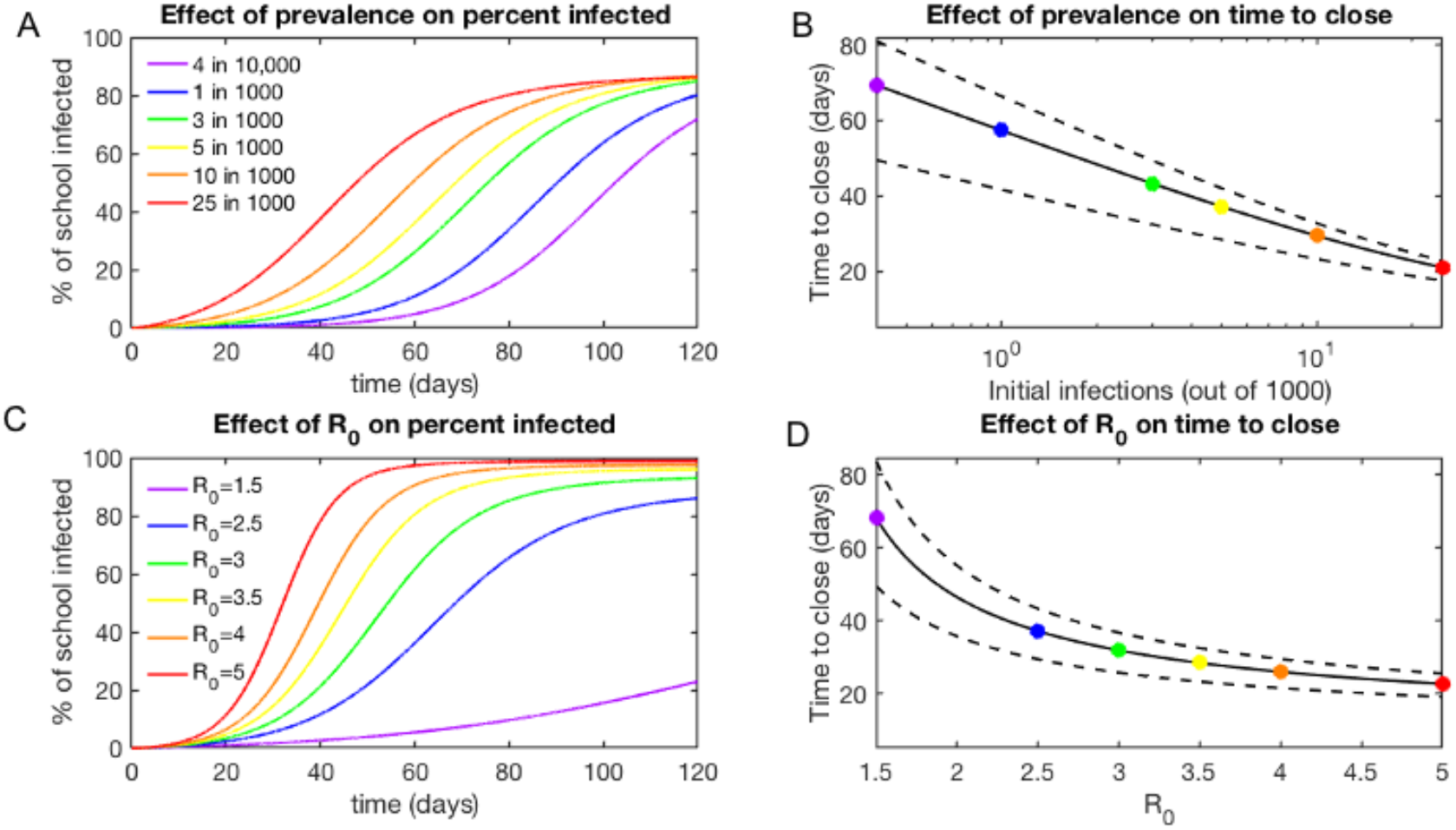
Clusters will develop quickly in schools upon reopening. A. Simulated time course of percent of school infected for initial confirmed prevalences ranging from 4 in 10,000 to 25 in 1000, demonstrating that a school reopening at low but non-zero prevalence of disease is simply delayed in its epidemic compared to schools with a higher initial disease prevalence. R_0_ = 2.5 for all projections. B. The effect of initial disease prevalence on the expected time to close, indicating that most schools in this regime will close between 20 and 60 days after opening. Upper and lower bounds reflect R_0_ bounds of 3.5 and 2.2 (See Methods: Estimation of School R_0_). C. Simulated time course of percent of school infected for reproductive numbers ranging from 1.5 to 5, demonstrating the speed at which high R_0_ s can lead to widespread infection. Initial prevalence is 5 in 1000 for all projections. D. The effect of Ro on the expected time to close, indicating again that most schools in this transmission regime will close between 20 and 60 days after opening. Upper and lower bounds reflect prevalence of 3 in 1000 and 10 in 1000 (See Methods: Estimation of average prevalence).

As of this writing, significant portions of the country are experiencing prevalences of COVID-19 in the range of 10 in 1000, and it is worth noting that schools that reopen in these ranges of prevalence will close quickly. In keeping with this, data on tracking websites, such as the Tableau public COVID-19 tracker for schools (*COVID-19 Reported Cases at U.S. Schools and Campuses*, 2020), reports over 11000 cases at 3000 schools across the country as of September 17^th^, 2020. In some regions, multiple schools have closed already, within days of reopening (Hanau, 2020). It is sobering to note that this is likely to be repeated on a much larger scale across the country in the coming weeks.

To determine the effect of transmission within the school on the rate of cluster growth and downstream consequences, we also examined the kinetics of cluster growth while varying the R_0_ across a reasonable range to be expected for schools with varying transmission reduction measures (Figure 1B). In this graph, the blue line marks an R_0_ of 2.5, a reasonable estimate for schools that have some measures in place to limit transmission. Schools reopening with an R_0_ =2.5 can expect to remain open for 40 days on average, and sooner if their threshold for detected cases is lower or if staff illnesses and high quarantine rates force earlier closure. Higher rates of transmission are certainly plausible for this disease in close quarters and will result in dramatically worse outcomes.

### Detected cases will form the tip of the iceberg

Despite the formation of clusters, in schools where systematic surveillance testing is not taking place, cases may be expected to mount undetected. Our modeling suggests that when clusters form in schools, the substantial contribution of asymptomatic and presymptomatic cases to transmission in the pediatric setting will create a discordance between the number of detected cases and the true scope of the outbreak (Figure 2A).

**Figure 2:**
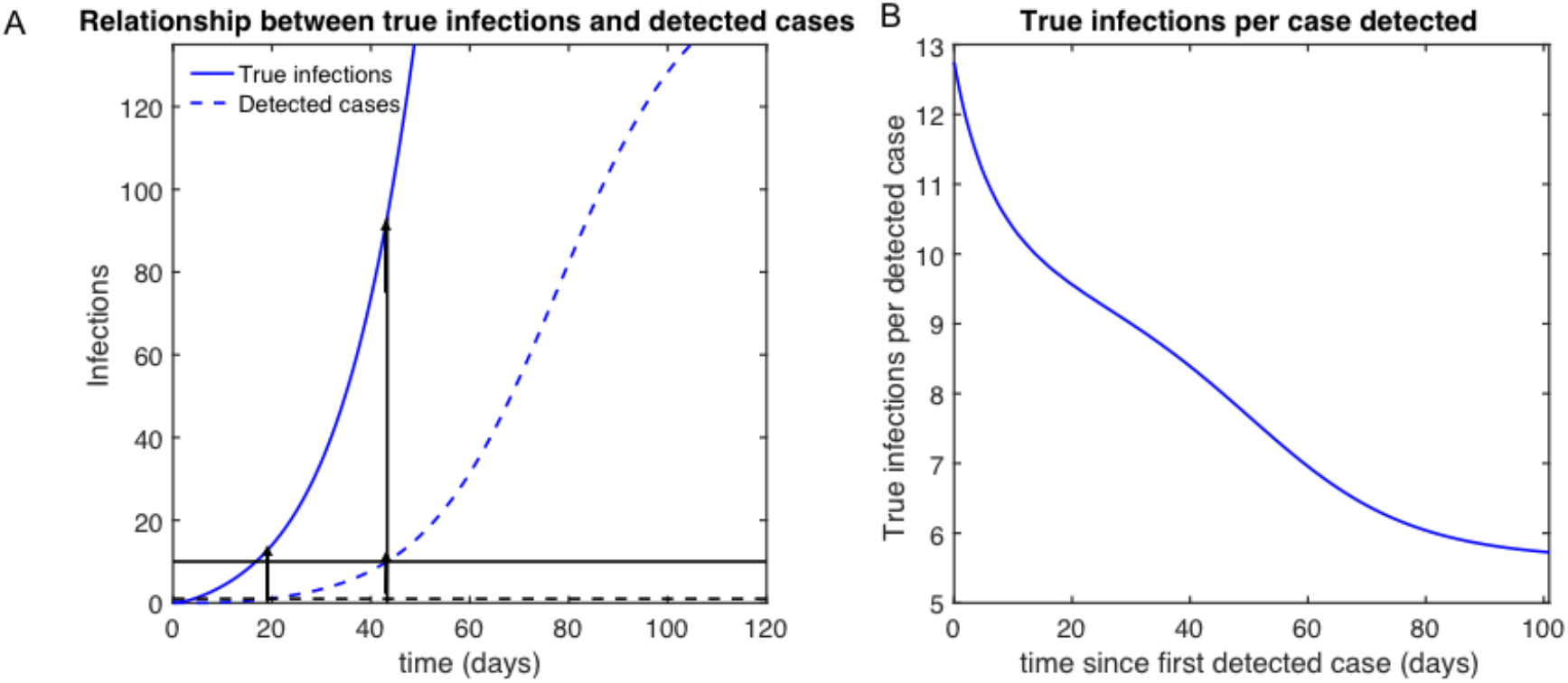
Detected cases will form the tip of the iceberg. A. Example of time course of infections (blue solid line) compared to time course of detected cases (blue dashed line), demonstrating a significant delay and under counting of reported student cases. Horizontal dashed black line represents the threshold for first detected case, and the horizontal black solid line represents the threshold for time to close of 10 detected cases in a school of 1000. Vertical arrows indicate the points on the true infection curve corresponding to the time of detection of the first case and school closure. B. Number of true infections per case detected over time, indicating that early in the outbreak due to significant delays in being tested or receiving a diagnosis, true infections can be 15x greater than detected cases.

This discordance is greatest at the beginning of the outbreak (Figure 2B), resulting in a 5 to 15-fold larger number of infections than detected cases at any time during the outbreak. As this ratio is highest at the beginning of the outbreak, it is precisely those schools that set the lowest thresholds (for the number of cases triggering closure) that will have the least understanding of the true scope of the outbreak at the time of closure. This finding has implications for controlling the secondary spread of COVID-19 through the community, as schools that shut down with only a few detected cases may still be seeding their communities with a significantly larger initial number of cases than is appreciated at the time of school closure.

### Schools will have limited room to maneuver in reopening safely

A recurrent theme in the reopening of the country has been the identification of conditions that permit safe reopening of schools. At the state level, a number of Departments of Education have left the identification of these conditions to individual schools and local health authorities (Florida Department of Education, 2020; Georgia Department of Public Health, 2020; Texas Education Agency, 2020). From an epidemiological perspective, there is value to examining the consequences of reopening in the face of varying rates of case prevalence and transmission. It is particularly noteworthy that schools may in fact not be guaranteed to act rationally in controlling the spread of COVID-19. An example of this would be schools crowding students into hallways without masks, then closing for two days to clean surfaces after an outbreak is reported (Andone and Johnston, 2020). With this in mind, we examined the impact of varying the local prevalence of COVID-19 as well as the school reproductive number (R_0_) on the time to the first detected case and the time to closing. We varied the transmission rate from 1 to 5 (the originally reported R_0_ from Wuhan at the beginning of the outbreak), and the prevalence from 0.1 to 10 out of 1000 (with the national average prevalence of this writing being 5). Across a wide range of conditions, the time to closing was 40 days or less (Figure 3B). Looking more closely at the conditions required for safe reopening of schools within the United States (Figure S2), our modeling suggests that an in-school R_0_ of less than 1.5 and a prevalence of less than 1 in 1000 is required for schools to remain open for 100 days or more.

**Figure 3:**
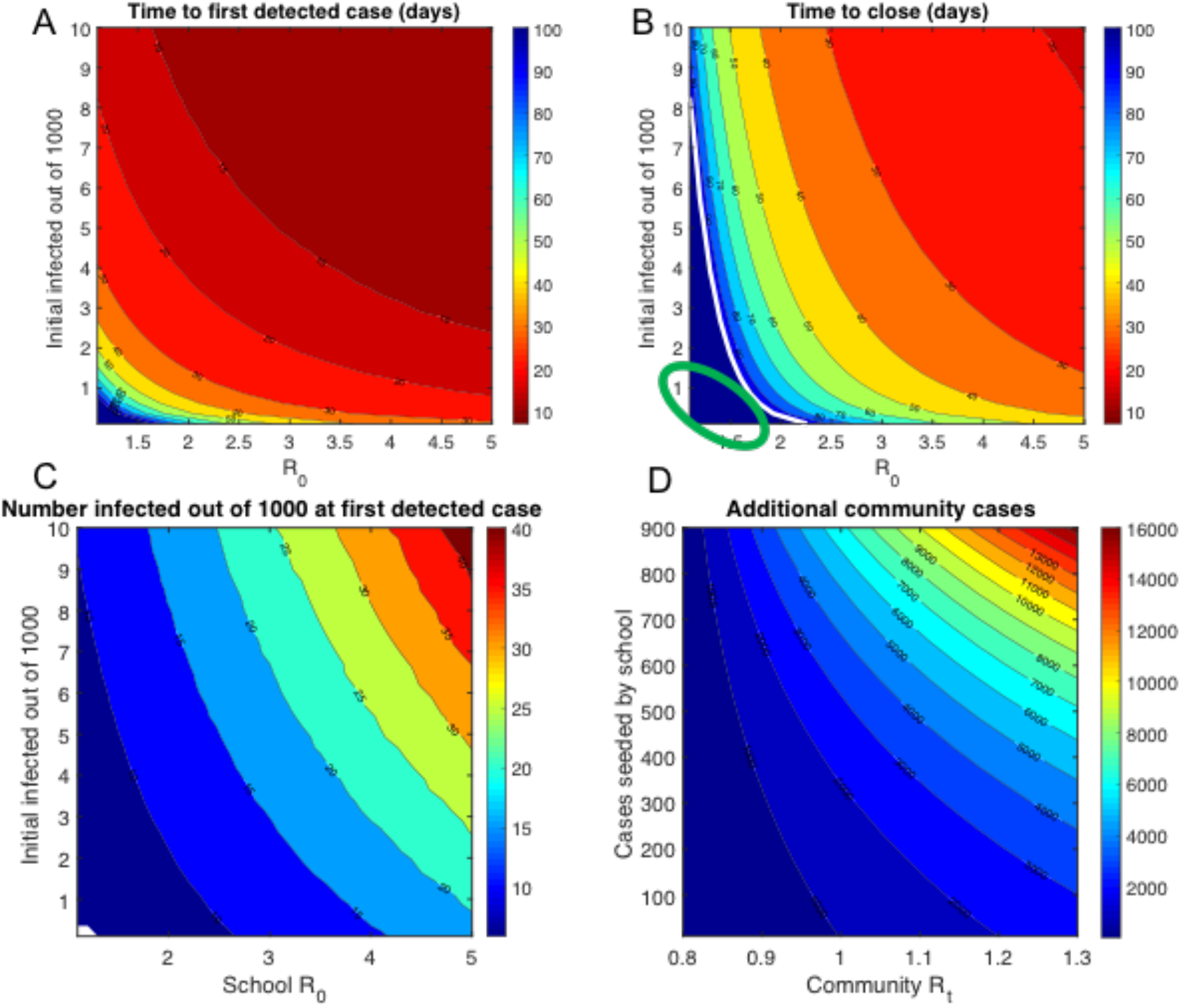
Schools have limited room to maneuver in reopening safely. A. Time to first detected case as a function of initial prevalence and school R_0_. Under many conditions, the time to detect the first case in a school is less than 20 days. B. Time to school closure as a function of initial prevalence and school R_0_. Most scenarios indicate school shutting down within 40 days. For a small subset of schools with R_0_ below 1.5 and initial prevalence below 1 in 1000, it may be possible to keep schools open for greater than 100 days. C. Number infected at first detected case as a function of initial prevalence and school R_0_, indicating even at the first detected case, the true number of infections at that time could be anywhere between 10 and 60. D. Additional community cases in the next 100 days as a function of infections seeded by school and a community’s R_0_, indicating the extreme risk for significant secondary infections into the community.

To provide more context for the outcomes that can be expected, we explored a few case studies of simulated K-12 schools, each with 1000 students (Table 1) with a range of different infection prevalences and reproductive numbers. While there is a wide range of potential outcomes possible, most scenarios result in schools reclosing within 60 days. The one noteworthy exception to this was the case of small schools that have low prevalence of the disease and a low in-school R_0_, which would be expected to remain open for approximately four months before having to close again.

**Table 1:**
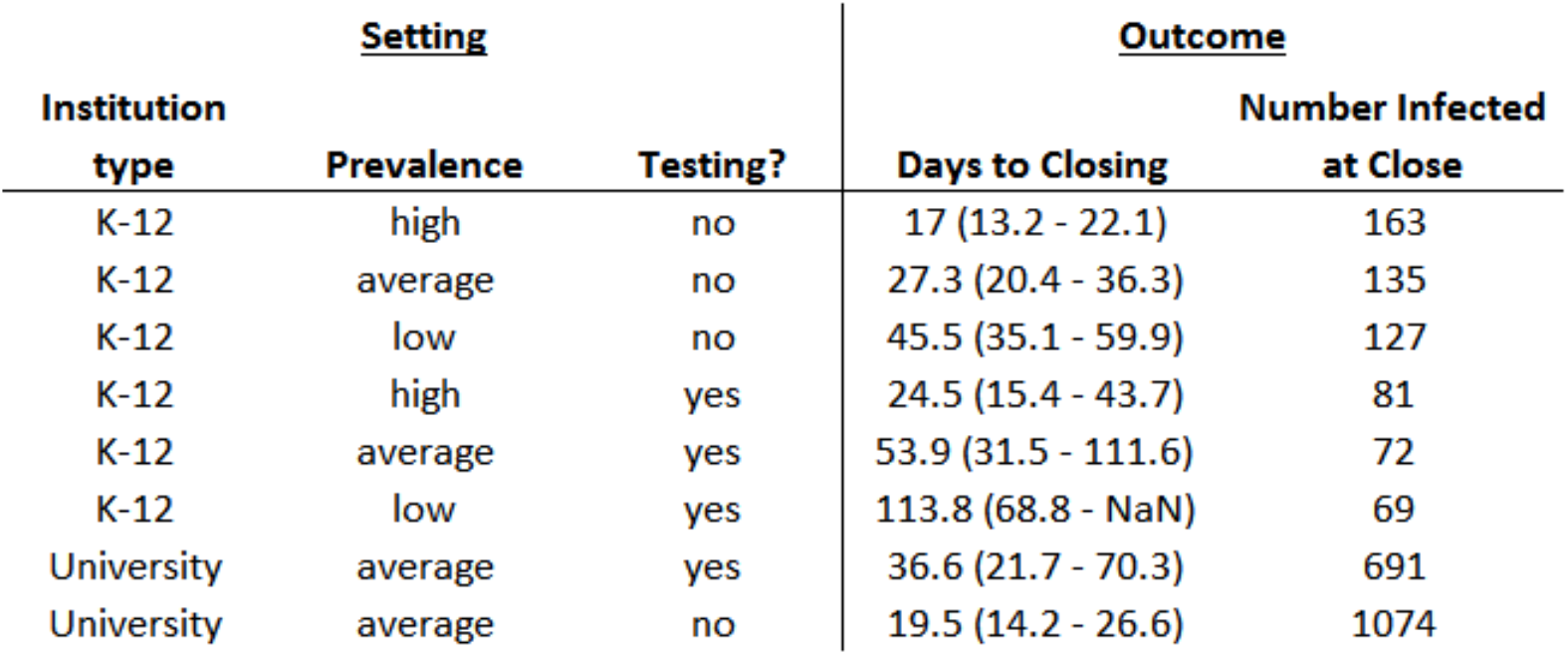
Example scenarios for a range of hypothetical settings corresponding to currently relevant rates of prevalence and transmission. In the absence of widespread testing to reduce transmission and very low prevalence, most schools will have to shut down within 60 days. The prevalence conditions correspond to high: 25 per 1000 (15,50 lower and upper bounds), average: 5 per 1000 (3 and 10 lower and upper bounds), low: 0.5 per 1000 (0.3 and 1 as lower and upper bounds). For schools without testing, we assumed an R_0_ = 3 (lower and upper bounds of 2.5 and 3.5), and for schools with rapid widespread testing we assumed an R_0_ = 1.5 (lower and upper bounds of 1.2 and 1.8).

As a sanity check, we also examined model predictions for the simulated scenario that would correspond to that of prestigious state schools (universities with 20,000 students drawn from across the country). Our model predicts the rapid emergence of large clusters of COVID-19 in this setting, consistent with recent events. As of this writing, multiple such colleges have seen the emergence of large clusters (Fowler, 2020; Levenson, 2020; The New York Times, 2020c) with over 50,000 cases in one month contributed by outbreaks at over 1000 colleges. It remains to be seen how much additional spread is contributed beyond the boundaries of these colleges as a result of reopening.

### Clusters originating in schools will seed further transmission

Once a cluster is initiated within a school population, it is unlikely that closing the school will end the transmission chains. Robust contact tracing for populations in close quarters is challenging, and the record so far indicates that quarantine and contact tracing protocols have been implemented in a haphazard manner (NBC 10 Boston, 2020; Price, 2020; Sheets, 2020). Behaviorally, it is plausible that students may continue to socialize in their community after school closure, potentially motivated by factors such as boredom (Wolff *et al*., 2020). Thus, it is reasonable to assume that a cluster of cases, once initiated within a school, will continue to spread within the community. Parenthetically, this effect has already been observed in smaller counties with colleges (Snyder, 2020a). Even students who are behaving safely after school closure are still interacting with their household members, presumably without PPE, potentially creating small family clusters of the disease.

To get a sense of the magnitude of the impact on communities as a result of this secondary spread, we simulated the seeding of an outbreak within the community, using rates for community spread that are consistent with the R_t_s for the US at present (Systrom and Vladeck, 2020) (Figure 3D). Our analyses suggest that, unless measures are taken to limit spread from schools that have seen clusters, community spread could result in a very substantial amplification of infections, far exceeding the original scale of the outbreak. Coupled with the limited understanding of the extent of disease spread in schools that are not testing systematically, the extended spread of the disease through the community as a whole (even at low R_0_s within the community) creates a dramatic and underappreciated externality as a result of decisions to reopen schools-one that was not part of the national conversation at the time that schools elected to reopen.

## Discussion

The question of what will happen when schools reopen across the United States, in the face of sustained community transmission of SARS-CoV-2, is a pressing one. As the science around children’s susceptibility to SARS-CoV-2 and the mechanism of transmission of COVID-19 has evolved, a simple question to ask is: given what we know now, what steps can we take to keep our communities safe as schools reopen?

The work described in this paper seeks to answer that question, using an epidemiological modeling framework that is updated with the current state of knowledge. We varied in-school R_0_ and local prevalence across plausible ranges and modeled both the immediate downstream and the longer-term consequences of school reopening. While we did not explicitly model the impact of specific interventions (such as masks, testing or podding) on controlling disease spread, our modeling accounted for these interventions implicitly, as reflected by the range of R_0_ values tested.

The goal of our modeling was not divination (foreseeing the future) but connecting the decision to reopen in the face of different prevalences and R_0_ values to the downstream impact on the schools and communities as a whole. While we cannot predict the reactions of schools to the emergence of clusters, we can predict what will happen to disease spread within the school and the community at large when clusters have emerged.

Clusters will form quickly within schools upon reopening, based on our modeling performed across a range of prevalences that mirrors the situation in the country right now. This has already been observed with colleges, although to some extent college outbreaks have been portrayed as a result of individual failings of college students, rather than as a structural consequence of prolonged indoor contact among large numbers of people in environments where the virus continues to spread (see Supp. Table. S4 for further examples of indoor spread).

These in-school disease clusters may not be apparent at first. Given the higher proportion of asymptomatic cases among children, the detected cases will conceal a far larger burden of infection within the school setting. In examining the range of prevalence and transmission rates that allows for safe reopening, we find that there is a window where schools can reopen for up to 100 days without the emergence of a disease cluster exceeding a cumulative 1% of the student body with detected cases: with community prevalences below 1 in 1000 and in-school R_0_s below 1.5. As to whether schools will be able to meet these standards with the available safety measures is an open question. In reality, many schools may be unable to provide enough space, PPE, sanitization to meet the CDC’s guidelines. In the absence of surveillance testing, schools will rely on symptomatic isolation, which we and others have shown to be ineffective at controlling disease spread (Johnson *et al*., 2020). More worryingly, there has been no evidence-based assessment of the efficacy of the current CDC guidelines in preventing disease spread.

Without testing or contact tracing, disease clusters seeded at schools will continue to spread silently throughout our communities, even after school closure. In our modeling, we find that school clusters, once initiated, have the potential for seeding enormous transmission chains within their communities, even at the lower R_0_s associated with community spread. Given the high proportion of asymptomatic cases within children, and the current testing guidelines, these clusters may spread within the community for a considerable period of time before they are detected.

As a purely practical matter, our work has three implications for successful school reopening. First, it points to the importance of taking measures to estimate the in-school prevalence of infections as early as possible. Rapid and widespread testing in general plays a key role in reducing transmission as a number of model-based analyses have shown very effectively (Berger, Herkenhoff and Mongey, 2020; Giordano *et al*., 2020; Larremore *et al*., 2020; Taipale, Romer and Linnarsson, 2020). To this end, the approval of a rapid antigen-based test for SARS-CoV-2 holds great promise. Widespread deployment of surveillance testing into the community represents one means of preventing explosive growth of COVID-19 within communities. If this is not possible, schools should consider pegging their re-closing criteria to county-level disease prevalence and not the number of cases in the school.

Second, it is critical that guidelines for limiting indoor spread of the virus be updated by public health authorities in an evidence-based manner (Morawska *et al*., 2020), with a reassessment of the risks posed by possible aerosol and fecal-oral transmission. Beyond handwashing, mask wearing and physical distancing, there are a number of measures that schools can take to reduce R_0_ from its initial baseline-for example improved ventilation, widespread surveillance testing, contact tracing-based isolation and cohorting/podding to reduce time in the classroom (the hybrid model). Widespread surveillance testing and rigorous contact-tracing based isolation are not being broadly implemented in K-12 schools across the US, even though they have demonstrated their utility in other countries. However, many schools within the US have adopted some of these additional measures, in particular improving ventilation and reducing in-person time via the hybrid model. It is worth noting that the impact of the hybrid model is likely to be dependent on what students do on the days that they are not at school. In such a case, if students socialize on their days at home, or go to a daycare, the hybrid model may in fact not result in limiting transmission. Mapping the impact of specific measures to changes in the R_0_ is beyond the scope of this work, but a number of other papers have explored this question, using both retrospective analyses of contact-tracing data (Bielecki *et al*., 2020) and agent-based simulations (Alagoz *et al*., 2020; Cuevas, 2020). We urge school authorities to implement as many measures as possible to reduce the in-school R_0_, as this variable has a profound impact on the feasibility of school reopening. Mandating mask use, upgrading ventilation systems, cohorting students, requiring students to continue to social distance/stay home when not at school and implementing widespread surveillance testing (particularly if rapid, easily performed tests are available) are all interventions worth considering if feasible.

Finally, schools need to be conscious of the externalities that will be forced upon their communities that they live in as a result of decisions to reopen in an unsafe manner. To this extent, should schools choose to close again as the result of an outbreak, aggressive measures will need to be taken to limit the spread of disease in areas undergoing community transmission. Failure to take this step could lead to the newly-seeded school clusters fueling chains of transmission within their communities that continue for months. To this end, our work suggests the value of quarantining all students following school closure and implementing surveillance testing in communities that have had school closures, if possible.

Sustained community transmission of SARS-CoV-2 has prolonged the pandemic within the United States, forcing a set of hard choices at the community and individual levels within the country. As the long-term nature of the pandemic becomes apparent, the debate around whether or not children should return to school has been framed as a choice between the health risks of COVID-19 specifically for children and the educational and social benefits of a return to school for children. A number of arguments can be raised against this framing-given that children are taught in schools by (and in most cases, live at home with) adults, the cost-benefit of a return to school cannot be examined for them in isolation. Critically, if children’s return to school spikes a chain of transmission that percolates into their families and communities, then the debate is being framed as a false dual choice. It is possible for children’s return to school to create a situation where they compromise their health and the health of their families and communities, while also leading to school shutdowns and depriving them of the benefit of in-person learning.

While we recognize the clear benefits of in-person education, we urge school authorities across the country to reflect on the downstream consequences of reopening. It can readily be agreed that in-person learning is vital for children’s development in the long run. However, our work suggests that school reopenings should be done with careful consideration paid to COVID-19 prevalence and measures to limit the in-school R_0_ as much as possible. If not, school reopenings will spawn undetected disease clusters, leading to an inevitable return to remote learning, and a long shadow of disease that spreads through our communities in the months following school closure.

## Data Availability

New York Times case counts by U.S. county were referred to in this manuscript and guided decisions made to produce the results of the manuscript. No data was directly downloaded. Code is available at https://github.com/kej1993johnson/School_reopening

https://www.nytimes.com/interactive/2020/us/coronavirus-us-cases.html

## Acknowledgments

The authors thank the UT COVID-19 Modeling Consortium for their feedback and suggestions on this work.

## Supporting Information Captions

**Figure S1**. Estimated number of actual infections at first detected case under A. varying initial number of infected people (in a school of 1000) at a constant R_0_=2.5 (lower and upper bounds represent R_0_= 2.2 and 3.5) and B. varying school reproductive number at a constant initial prevalence of 5 in 1000 (lower and upper bounds represent initial prevalence of 3 In 1000 and 10 in 1000).

**Figure S2. Estimated percent of school quarantined** at any time after school opening under A. varying number of people infected at the start at a constant R_0_=2.5 and B. varying school reproductive number at a constant initial prevalence of 5 in 1000.

