## Supplementary Information for "In the long shadow of our best intentions: model-based assessment of the consequences of school reopening during the COVID-19 pandemic"

| Parameter | Symbol | Value | Reference |
|---|---|---|---|
| Basic reproductive number: number of individuals an infectious individual infects on average | $R_0$ | 2.5 individuals [2.2, 3.5] | (CDC, 2020a) |
| Daily reproduction rate | $\beta$ | Basic reproductive number divided by duration of infection ($\beta\gamma$) | Calculated from $R_0$ |
| Duration of time exposed but not detectable or infectious | $1/\alpha$ | 3 days | (Lauer *et al.*, 2020) |
| Duration of infection | $1/\gamma$ | 14 days | (He *et al.*, 2020) |
| Time from initial infection to seek testing if symptomatic | $\tau_{seektest}$ | 2.3 days (before symptoms) + 0.7 delay to get tested = 3 days | (He *et al.*, 2020) |
| Time to receive test result | $\tau_{testdelay}$ | 4 days | (Baum *et al.*, 2020) |

**Table S1. Model parameters and references for SEIR model**

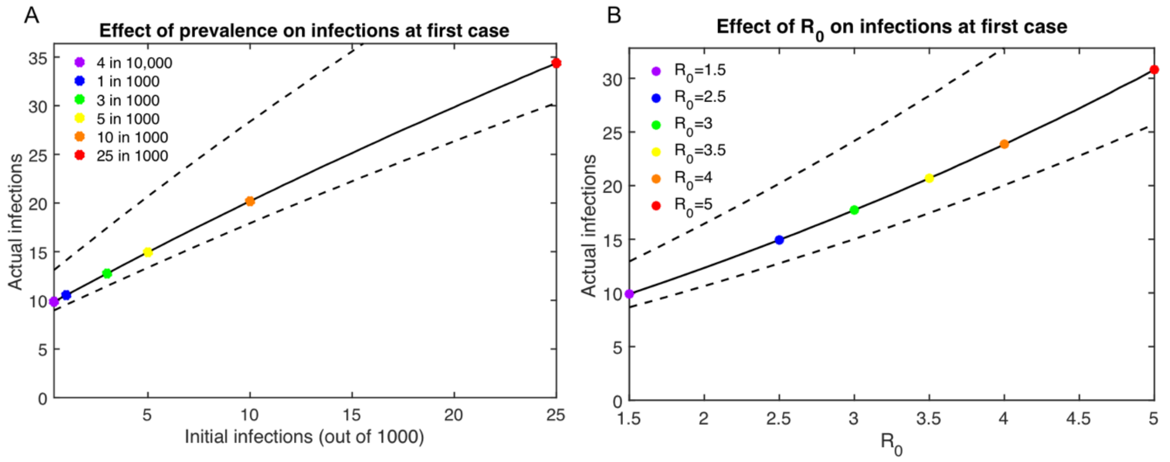


**Figure S1.** Estimated number of actual infections at first detected case under A. varying initial number of infected people (in a school of 1000) at a constant R0=2.5 (lower and upper bounds represent $R_0$= 2.2 and 3.5) and B. varying school reproductive number at a constant initial prevalence of 5 in 1000 (lower and upper bounds represent initial prevalence of 3 In 1000 and 10 in 1000).

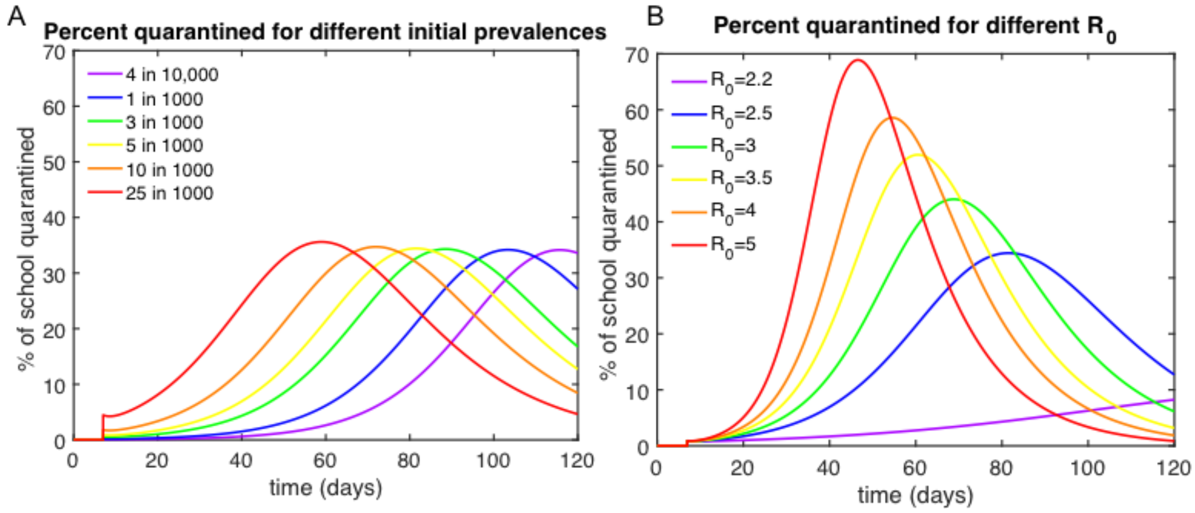


**Figure S2. Estimated percent of school quarantined** at any time after school opening under A. varying number of people infected at the start at a constant $R_0$=2.5 and B. varying school reproductive number at a constant initial prevalence of 5 in 1000.

| Location | NY Times cases per 100k in the last 7 days as of September 5th, 2020 (The New York Times, 2020) | Estimate of prevalence per 1000 |
|---|---|---|
| Lafayette County, Mississippi | 609 | 30.5 (18.27-60.9) |
| Baldwin County, Georgia | 370 | 18.5 (11.1- 37.0) |
| Dallas County, Texas | 121 | 6.05 (3.63- 12.1) |
| Union County, New Jersey | 24 | 1.2 (0.72- 2.4) |
| Cheshire County, New Hampshire | 11 | 0.55 (0.33 – 1.1) |
| United States | 88 | 4.4 (2.6 – 8.8) |

**Table S2. County prevalence values for different locations in the US**, assuming a 1 in 5 case reporting rate an infectious duration of 7 days. Upper and lower bounds of prevalence estimates are for a range of 1 in 10 and in 1 in 3 case reporting rate.

To estimate the prevalence per 1000 in a county, we take the cases per 100 thousand in the last 7 days from the New York Times website (The New York Times, 2020). For example, in Dallas County, Texas there were 121 cases per 100 thousand in the last 7 days. We assume those represent all active infections. Then we assume that on average 1 in 5 infections are detected, and so the true infection prevalence is 5 times greater, yielding 605 infections per 100 thousand. We divide by 100 to get cases per 1000. Thus it is expected that in a school of 1000, about 6 individuals will show up infected in the first week of school (with a range of 4 to 12 depending on case reporting rate).

| **Scenario** | **Estimate of $R_0$** | **Method of $R_0$ estimation** | **Reference** |
|---|---|---|---|
| Wuhan, China prior to SARS-CoV-2 detection and lockdown | 1.4- 5.7 | Model-based inference | (Majumder and Mandl, 2020) (1.4 -4)<br>(Sanche *et al.*, 2020) (5.7 (3.8-8.9)) |
| Diamond Princess Cruise Ship | 14.8 | Model-based inference | (Rocklöv, Sjödin and Wilder-Smith, 2020) |
| Early epidemic in Europe and the US | 4.0-7.1 | Model-based inference | (Ke *et al.*, 2020) |

**Table S3. $R_0$ of COVID in different settings.** In settings where large indoor gatherings were still taking place, or where social distancing protocols were difficult to implement, baseline $R_0$s for SARS-Cov-2 were considerably higher than the $R_0$s in states within the US, which are currently around 1.

| Scenario | Transmission reduction measures | Outbreak details | Reference |
|---|---|---|---|
| Israel school reopening | masks required and windows opened but then a heat wave prompted relaxation and masks were removed and windows closed | Potential superspreading: 154 students and 26 staff members infected | (Kershner and Belluck) |
| SUNY Campus August 2020 | social distancing measures in place but student parties still occurred | More than 500 students positive in the first two weeks out of a school of 6,000 (where only 30 initial infections would have been expected) | (Ross, 2020) |
| Controlled cohort study on an 11 hour flight | symptomatic individuals screened and removed, N95 masks provided and worn except for meals and restrooms | One woman expected to have been infected from sitting 3 rows away from asymptomatic patient | (Bae *et al.*, 2020) |
| Outbreak in indoor choir in March 2020 in Seattle | no masks but at half-capacity | 2.5 hour choir practice- 52 of the 60 choir members became ill from single index patient | (Hamner *et al.*, 2020) |
| Outbreak in air-conditioned restaurant in Guangzhou, China in late January 2020. | Tables 1 meter apart | 9 others (4 from family of infected individual, 3 members from additional family and 2 members from another family) | (Lu *et al.*, 2020) |
| Outbreak in call center in South Korea in March 2020 | None- crowded with lots of talking | 94 employees on a the 11$^{th}$ floor of the building, 216 were infected by a single index case | (Park *et al.*, 2020) |

**Table S4. Documented indoor outbreaks with different levels of transmission reduction behaviors**
